# Surrogate virus neutralisation test based on nanoluciferase-tagged antigens to quantify inhibitory antibodies against SARS-CoV-2 and characterise Omicron-specific reactivity

**DOI:** 10.1101/2023.10.10.23296792

**Authors:** M Schoefbaenker, R Neddermeyer, T Guenther, MM Mueller, ML Romberg, N Classen, MT Hennies, ER Hrincius, S Ludwig, JE Kuehn, EU Lorentzen

## Abstract

Virus-specific antibodies are important determinants of protective immunity against severe acute respiratory syndrome coronavirus 2 (SARS-CoV-2). While regarded as the gold standard for detecting functional antibodies, conventional virus neutralisation tests (VNT) or pseudotyped virus neutralisation tests (pVNT) require biosafety level 2 or 3 facilities. Alternatively, the virus-free surrogate virus neutralisation test (sVNT) quantifies inhibitory antibodies that prevent the spike protein from binding to its receptor, human angiotensin-converting enzyme 2 (hACE2).

We evaluated secreted nanoluciferase (NLuc)-tagged spike (S) protein fragments as diagnostic antigens in the sVNT in the framework of a vaccination study. First, spike fragments of different lengths were tested for their suitability as diagnostic antigens in a capture enzyme immunoassay (EIA) using unprocessed culture supernatants of transfected cells, identifying the receptor binding domain (RBD) of S as the optimal construct. The sensitivity of the in-house sVNT relying on the NLuc-labelled RBD equalled or surpassed a commercial sVNT (*cPass*, GenScript Diagnostics) and an in-house pVNT four weeks after the first vaccination (98% vs. 94% and 72%, respectively), reaching 100% in all assays four weeks after the second and third vaccinations. Additionally, serum reactivity with spike constructs of Omicron BA.1 was tested. Compared with a capture EIA, the in-house sVNT and pVNT displayed superior discrimination between wild-type- and variant-specific reactivity of sera. Differences in reactivity were most pronounced after the first and second vaccinations, whereas the third vaccination resulted in robust, cross-reactive detection of Omicron constructs.

In conclusion, assays utilising NLuc-labelled protein fragments permit the quantification and functional assessment of SARS-CoV-2-specific antibodies and the detection of variant-specific differences in reactivity. Potential applications include monitoring therapy and vaccine efficacy and follow-up of prolonged disease courses in high-risk groups. Designed as straightforward, highly flexible modular systems, these tests can be readily adapted to further emerging viral variants.

## INTRODUCTION

Since the WHO declared coronavirus disease 2019 (COVID-19) a pandemic in 2020, vaccines based on the original Wuhan-Hu-1 strain of SARS-CoV-2 and its descendants, specifically Omicron BA.1 and BA.4/5, have proven to be effective in controlling the disease [1]. These vaccines mainly target the viral spike (S) surface glycoprotein as the predominant mediator of immunity [2, 3]. Immune responses following infection, vaccination or both (known as hybrid immunity) are under thorough investigation to determine the extent and duration of protection against emerging virus variants. Studies on pre-Omicron variants have shown that hybrid immunity provides optimal and long-lasting protection against severe disease, an important finding that is already influencing current vaccination schemes [4]. Furthermore, enhanced immune responses have been observed following vaccination post-infection, such as an increase in antibodies directed against the more conserved domain 2 of the SARS-CoV-2 spike protein (S2), which have been identified and verified in various variants, including Omicron B.1.1.529 and BA.5 [5]. While immunity mediated by B and T cells appears to persist, with an even greater effect noted in hybrid immunity, antibody levels gradually decrease over time. However, booster vaccinations have shown their efficacy in inducing broad humoral immunity against Omicron variants BA.1, BA.2 and BA.5 [6]. Conversely, prior infection or vaccination may impede neutralising immune responses to subsequent viral variants such as B.1.1.529, BA.4 or BA.5 [7, 8]. In addition to monitoring population-level developments in (hybrid) immunity and immune responses in high-risk patient groups, it will be crucial to project the efficacy of vaccines developed to counter evolving immune escape variants in order to guide future vaccination strategies.

Hence, serologic characterisation of neutralising antibodies has increasingly gained importance, particularly regarding emerging escape variants. To this end, standard serologic tests that detect SARS-CoV-2-specific antibodies irrespective of their neutralising capacity, such as enzyme-linked or chemiluminescent immunoassays (ELISA, CLIA), have to be complemented by functional virus neutralisation assays. The majority of neutralising antibodies in convalescent sera target epitopes within the receptor binding domain (RBD) of the viral spike protein domain 1 (S1), which mediates attachment to its cellular receptor, human angiotensin-converting enzyme 2 (hACE2), and thus, host cell entry [9–13]. As the RBD displays sufficient variability to discriminate between different human coronaviruses and simultaneously high specificity when used as a diagnostic antigen, it has become the target of choice in serologic assays [14, 15]. In addition, the N-terminal domain (NTD) of S1 has been shown to harbour neutralising epitopes recognised by convalescent and vaccinee sera [16–18]. Viral variants frequently evade neutralising immune responses or therapeutic monoclonal antibodies through mutations within immunodominant epitopes such as E484 of the RBD, with Omicron and its subvariants accumulating more than 30 mutations within spike [11, 19–23]. Whereas minimal protective antibody titres have not yet been defined, neutralising antibodies are regarded to correlate with protection from severe disease, rendering them suitable surrogate markers for immunity and vaccine efficacy studies [24].

While conventional virus neutralisation tests (cVNT) such as plaque reduction neutralisation tests (PRNT) are considered the gold standard, they require successful and time-consuming virus isolation and biosafety level (BSL) 3 containment, which impedes many diagnostic laboratories. As an alternative, vector-based pseudotyped virus neutralisation tests (pVNTs) have been developed enabling faster testing within standard BSL-2 facilities, however, as in-house tests they still lack standardisation. To facilitate the assessment of neutralising antibodies, surrogate virus neutralisation tests (sVNT) have been introduced to the market that can be conducted in a standardised format under clinical routine BSL-2 laboratory conditions within only a few hours [25–27]. These assays rely on the inhibition of SARS-CoV-2 RBD binding to hACE2 and have been shown to correlate well with cVNTs and various SARS-CoV-2 S protein-based pVNT assays, rendering them valuable tools for high-throughput functional antibody screening [25–33]. In the wake of the first primary Omicron infections, commercial antibody assays targeting the S protein based on the ancestral Wuhan-Hu-1 strain of SARS-CoV-2, including sVNTs, have revealed a decline in sensitivity, warranting adaptation of diagnostic antigens to the currently prevailing variants [34].

To investigate the influence of variant-specific mutations on the reactivity of vaccinee sera and to differentiate between different immunoglobulin classes without handling infectious virus particles, we developed virus-free assays applying S protein fragments of varied lengths, including the RBD and NTD (**Fig. 1**; **Table 1A**) linked to nanoluciferase (*NanoLuc*, NLuc, cf. Materials and Methods) as bioluminescent reporter-tagged antigens. The nanoluciferase reporter system has been demonstrated to be appropriate for immunoassays such as the fluid-phase luciferase immune precipitation (LIPS) assay [35]. In our assessment of a cohort of vaccinated individuals, our in-house sVNT and immunoglobulin capture enzyme immunoassay (EIA) complemented routine serologic tests convincingly, displaying high levels of sensitivity, specificity, and reproducibility. The accuracy of the findings was confirmed through comparison with commercial serologic assays, as well as an in-house VSV-based pVNT (VSV-pVNT) employing a vesicular stomatitis virus (VSV) bearing the SARS-CoV-2 S protein. The sVNT and capture EIA are highly adaptable and simple assays that allow rapid analysis of variant-specific humoral immune responses.

**Figure 1:**
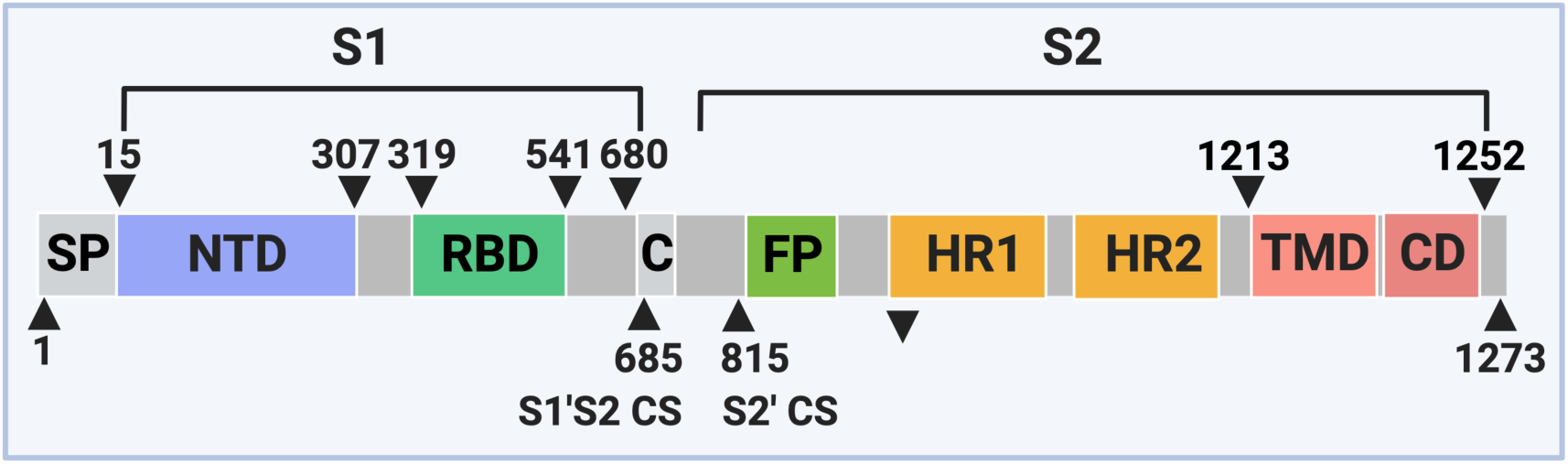
SARS-CoV-2 spike protein primary structure. S1: spike protein domain 1, S2: spike protein domain 2, SP: signal peptide, NTD: N-terminal domain, RBD: receptor binding domain, C/CS: cleavage site, FP: fusion peptide, HR: heptad repeat, TMD: transmembrane domain, CD: cytodomain. The illustration was created with BioRender.com.

## MATERIALS AND METHODS

### Vaccinee sera

Serum samples were collected within the framework of a study approved by the ethics committee of the Aerztekammer Westfalen-Lippe, Muenster, Germany, and the University of Muenster (2021-039-f-S) regarding the *Characterisation of the Humoral Immune Response to SARS-CoV-2 in Vaccinees*. Participants received either *Comirnaty* (BNT162b2, BioNTech/Pfizer) or *Spikevax* (mRNA-1273, Moderna) mRNA vaccines based on wild-type Wuhan-derived S protein sequences. Samples were obtained at baseline before immunisation (t0) and 4 weeks post first vaccine dose (t1), 4 weeks (t2), 3 months (t3) and 6 months after the second vaccine dose (t4), and 4 weeks after (t5) the third vaccine dose, respectively. The sampling period ranged from January 2021 to January 2022.

### Routine serology

Systematically, IgG antibodies specifically targeting the receptor binding domain (RBD) of the SARS-CoV-2 S protein (anti-RBD IgG) were quantified using the commercial, CE/IVD certified chemiluminescence microparticle immunoassay (CMIA) *SARS-CoV-2 IgG II Quant* on an *Architect* platform (Abbott Diagnostics, Wiesbaden, Germany). Seropositivity was indicated by values greater than or equal to 50.0 arbitrary units (AU)/mL, where 0.142 Abbott AU/mL corresponds to 1 binding antibody unit (BAU) as defined by the WHO. Abbott claims a sensitivity of up to 99.37% depending on the day post-symptom onset and a specificity of 99.55%. Accordingly, the presence of IgG antibodies against the nucleocapsid (N) protein (anti-N IgG) of SARS-CoV-2 was qualitatively determined using the CMIA *Abbott SARS-CoV-2 IgG* in vaccinees to detect any potential infection (sensitivity 100%, specificity 99.63%). In addition, the *recomLine SARS-CoV-2 IgG CE/IVD* (Mikrogen, Neuried, Germany) commercial line blotting assay was applied to assess sera with equivocal anti-N IgG levels at t0. The test detects IgG directed towards the SARS-CoV-2 RBD, S1 and N antigens semi-quantitatively using a line blot format. All tests were conducted according to the manufacturer’s instructions.

The commercial, CE/IVD-certified *cPass SARS-CoV-2 Neutralization Antibody Detection Kit* (GenScript Biotech, Leiden, The Netherlands) was employed to semiquantitatively evaluate the neutralising activity of sera, using sequences derived from the Wuhan strain. This assay measures the degree to which antibodies inhibit the binding of the RBD to the hACE2 receptor, using RBD-HRP (horse-radish peroxidase) conjugates in a blocking ELISA format and is considered one of the most accurate sVNTs available [25, 27, 31]. Following the manufacturer’s manual, serum samples were tested in technical duplicates, with a final dilution of 1:20. The percentage of inhibition was calculated as (1 - OD value of the sample/OD value of the negative control) × 100%. Values below the cut-off threshold of 30% are indicative of a negative result; values equal to or exceeding the cut-off indicate the presence of SARS-CoV-2 neutralising antibodies.

### In-house assays

#### Pseudovirus-based virus neutralisation test

The in-house VSV-pVNT utilised a vesicular stomatitis virus (VSV) carrying the SARS-CoV-2 S protein. To enhance trans-complementation through the transiently expressed S protein in the VSV-pVNT, we employed site-directed mutagenesis to introduce a 21 amino acids (aa) long C-terminal deletion into the expression vector pCG1-SARS-2-S [10] that comprises the wildtype (wt) S protein aa sequence (GenBank ID NC_045512.2) [1]. The plasmid pCG1-SARS-2-S was cleaved with SalI and BsaBI. Subsequently, the product of primers S BsaB1 forward (fwd) and S Delta1253 backward (bwd) amplified from plasmid pCG1-SARS-2-S was inserted into the vector by *In-Fusion* cloning (Clontech/Takara Bio, Mountain View, CA, USA). The resulting construct was designated pCG1-SARS-2-S-Delta1253. The S protein with the Omicron BA.1-specific amino acid sequence and the C-terminal 21 aa deletion was expressed from vector pcDNA3.1 SARS-2 Omicron, containing a synthetic S-insert derived from BA.1 (Thermo Fisher Scientific, Schwerte, Germany).

SARS-CoV-2 neutralisation was analysed using the VSV-pVNT system (VSV ΔG/GFP-Luc + SARS-CoV-2 S protein variants) as described by [36]. The pseudotyped virus was generated according to the method outlined by [37]. The sera were diluted (1:20) and preincubated with the pseudotyped virus at 37°C/5% CO_2_ for 1 h (hour). Subsequently, Vero E6 cells were infected with the pseudotyped virus-sera mixture for 1 h at 37°C/5% CO_2_ (with a final multiplicity of infection (MOI) of 0.01). After 16 hours post-infection (h.p.i.), the number of GFP-positive cells was quantified with the *Celigo* Image Cytometer (Nexcelom/Perkin Elmer). Four technical replicates were measured, from which the average was calculated. A pool of 17 sera containing high levels of neutralising antibodies against S, collected at t2, was used as a positive control (PSP). Additionally, a corresponding pool of sera collected from the same patients at t0 served as a negative control (NSP). The degree of neutralisation was calculated as the reduction of the GFP signal (%) = (1 - GFP signal of the treated sample/GFP signal of the untreated sample) x 100%.

#### Cloning of RBD- and NTD-fragments of the SARS-CoV-2 S protein

Properties of plasmids expressing secreted S fragments are listed in **Table 1A**. The expression plasmid pEN-secNL-RBD containing an N-terminal *NanoLuc* (NLuc) luciferase tag (Promega, Walldorf, Germany) was generated by inserting amplified products comprising the secreted form of NLuc (secNL) and the RBD using two-fragment *In-Fusion* cloning into the vector pEGFP-N1 (Clontech), which had been opened with NheI and NotI. SecNL including the IL6 signal peptide was amplified from the plasmid pNL 1.3 (Promega) using primers secNL fwd and secNL-BamHI bwd. The RBD fragment was amplified from plasmid pCG1-SARS-2-S with primers RBD-BamHI fwd and RBD-NotI bwd (**Table 1B**). The expression plasmids pEN-secNL-S15-307, pEN-secNL-S15-541 and pEN-secNL-S15-680 were generated by inserting PCR fragments amplified from plasmid pCG1-SARS-2-S with the primer S15-BamHI fwd and primers S307-NotI bwd, RBD-NotI bwd and S680-NotI bwd (**Table 1B**), respectively, into pEN-secNL-RBD opened with BamHI and NotI. Expression plasmid pEN-secNL-RBD Omicron was obtained by inserting a fragment amplified with primers RBD BamHI_2 fwd and RBD NotI bwd from the vector pcDNA3.1 SARS-2 Omicron into pEN-secNL-RBD, which was opened using BamHI and NotI. The expression plasmid pEN-secNL-spike-CSdel-PPmut was created by inserting the product of primers S15-BamHI fwd and S CSdel-PPmut bwd amplified from the vector pCAspike-CSdeleted-PPmutation (GenBank ID MT380725.1) [15] as target sequence (**Table 1B**) into plasmid pEN-secNL-RBD.

**Table 1A:**
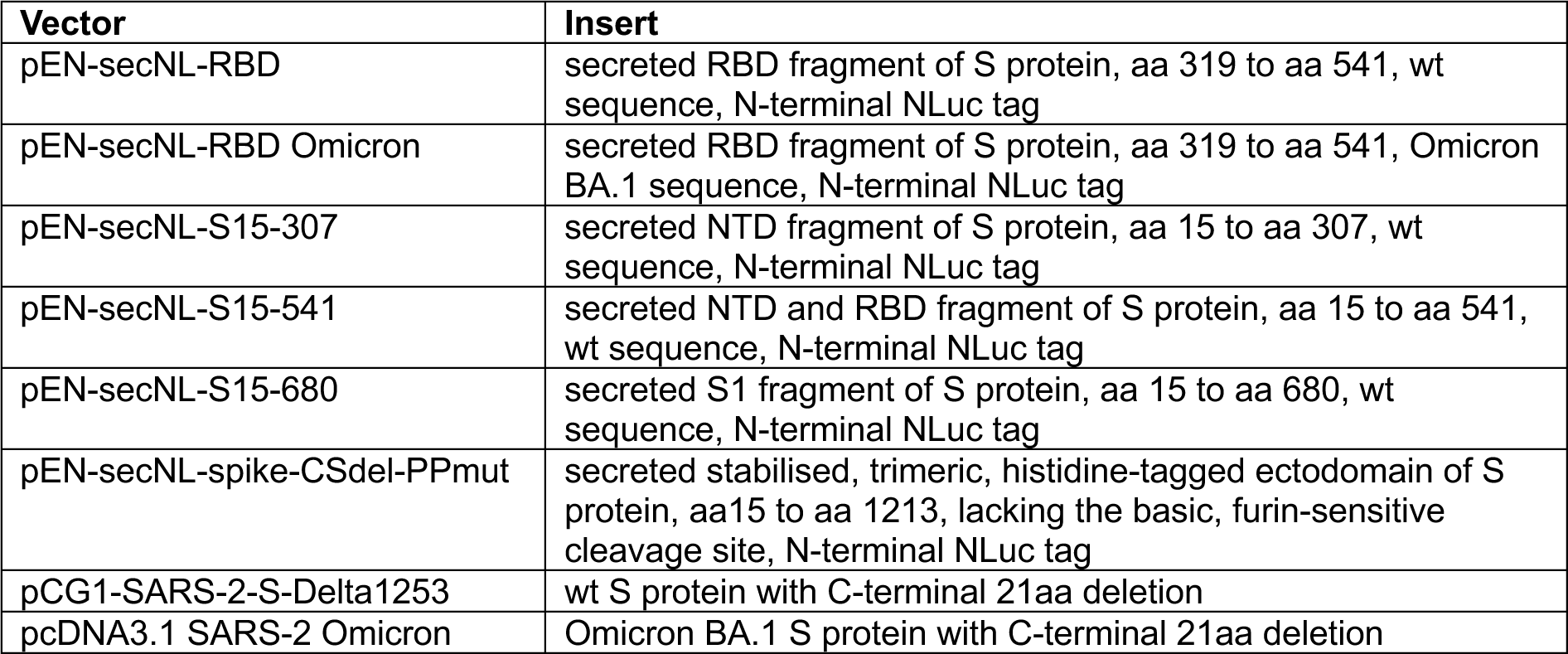
List of expression plasmids.

| Vector | Insert |
| --- | --- |
| pEN-secNL-RBD | secreted RBD fragment of S protein, aa 319 to aa 541, wt sequence, N-terminal NLuc tag |
| pEN-secNL-RBD Omicron | secreted RBD fragment of S protein, aa 319 to aa 541, Omicron BA.1 sequence, N-terminal NLuc tag |
| pEN-secNL-S15-307 | secreted NTD fragment of S protein, aa 15 to aa 307, wt sequence, N-terminal NLuc tag |
| pEN-secNL-S15-541 | secreted NTD and RBD fragment of S protein, aa 15 to aa 541, wt sequence, N-terminal NLuc tag |
| pEN-secNL-S15-680 | secreted S1 fragment of S protein, aa 15 to aa 680, wt sequence, N-terminal NLuc tag |
| pEN-secNL-spike-CSdel-PPmut | secreted stabilised, trimeric, histidine-tagged ectodomain of S protein, aa15 to aa 1213, lacking the basic, furin-sensitive cleavage site, N-terminal NLuc tag |
| pCG1-SARS-2-S-Delta1253 | wt S protein with C-terminal 21aa deletion |
| pcDNA3.1 SARS-2 Omicron | Omicron BA.1 S protein with C-terminal 21aa deletion |

**Table 1B:**
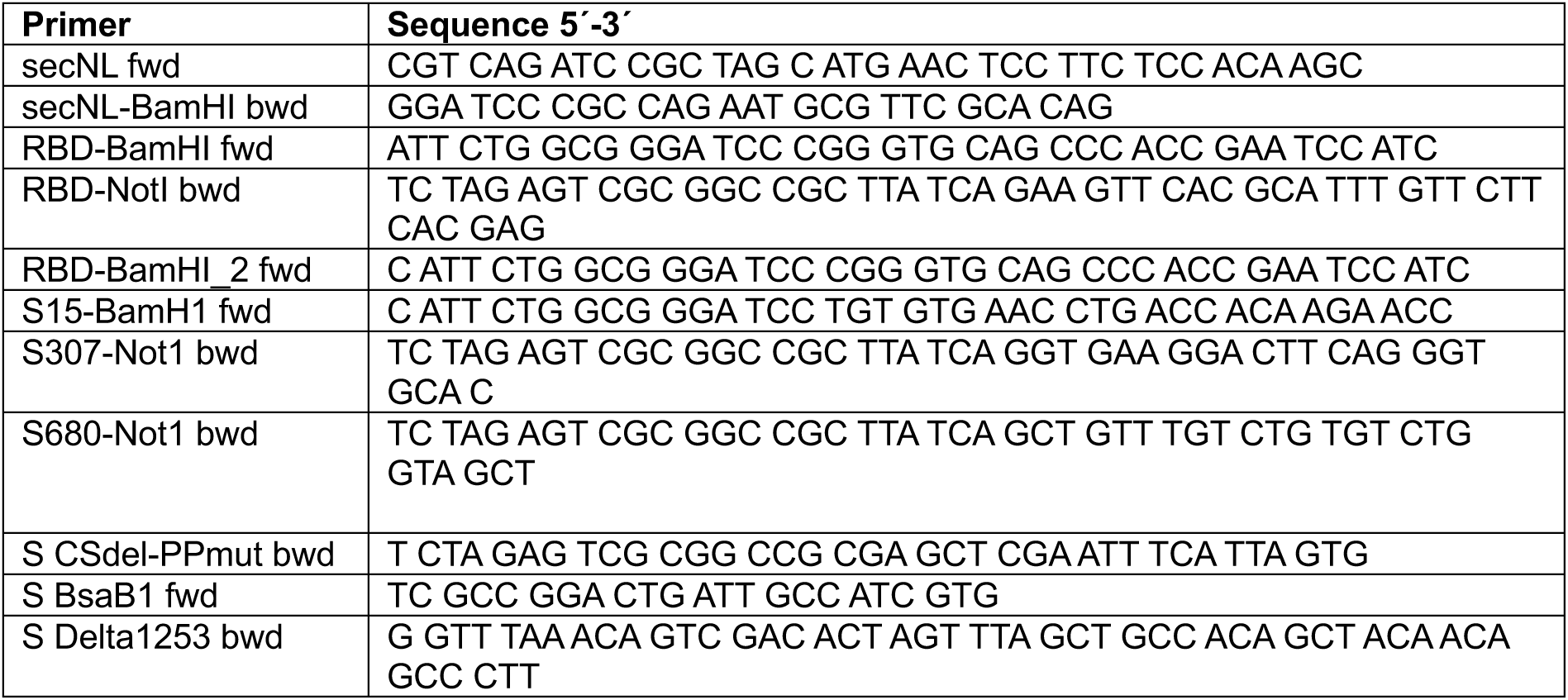
List of primers.

All plasmids were transfected into BHK cells cultured in 24-well plates using *lipofectamine 2000* (Thermo Fisher Scientific). Cell culture supernatants were harvested 24-48 h after transfection, and cell debris was removed by centrifugation (5 min at 3000 rpm). NLuc activity was determined in a *GloMax Explorer* reader (Promega) using *Nano-Glo* reagent (Promega). Finally, the samples were aliquoted and stored at -20°C until use.

#### Surrogate virus neutralisation test

The in-house surrogate virus neutralisation test (sVNT) was designed to measure the inhibition of the binding of secreted RBD-containing S protein fragments, which are N-terminally labelled with NLuc, to the hACE2 receptor in an ELISA format (**Fig. 2A**). Black high-binding 96-well plates (Greiner Bio-One, Frickenhausen, Germany) were coated with 200 ng/well of recombinant hexahistidine-tagged hACE2 (Sino Biological, Beijing, China, #10108-H08H) diluted in 100 µL carbonate buffer at 4°C for 16 h. The coated plates were washed four times using 300 µL/well TRIS-buffered saline (TBS)/0.1% Tween (TBS-T), blocked with 300 µL/well TBS-T/2% BSA (w/v) for 1.5 h at 37°C, and washed four times using TBS-T.

**Figure 2:**
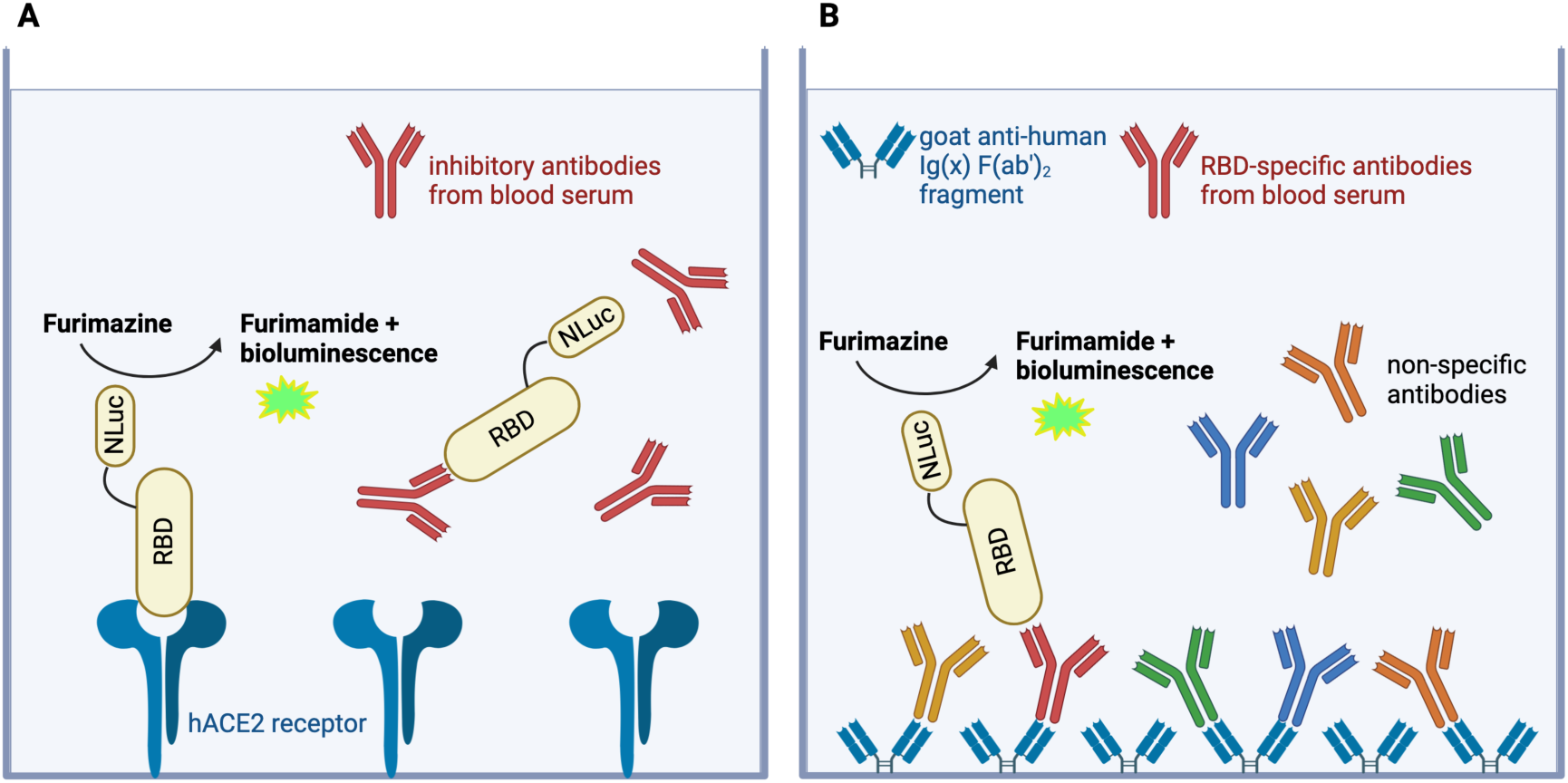
Schematic illustration of the in-house sVNT **(A)** and the immunoglobulin capture EIA **(B)** for characterisation of SARS-CoV-2-specific antibodies using RBD-NLuc fusion proteins from cell culture supernatants. Antibody-mediated inhibition of binding to hACE2 (A) and capture of RBD by specific antibodies (B) was quantified using nanoluciferase-mediated bioluminescence signals. NLuc: nanoluciferase; RBD: receptor binding domain; hACE2: human angiotensin-converting enzyme 2. The illustration was created with BioRender.com.

Subsequently, 110 µL of serum, which was diluted 1:20 in TBS-T and contained 5.5×10^6^ relative light units (rlu) secNLuc-RBD, underwent incubation for 30 min at 37°C. Following this, 100 µL/well were transferred to hACE2-coated 96-well microtiter plates and kept at 37°C for 1 h. The plates were washed with 300 µL/well TBS-T four times prior to the addition of 100 µL *Nano-Glo* reagent/well. Next, the resulting NLuc activity was quantified. Luciferase activity in the absence of human serum and SARS-CoV-2 antibody negative (NSP) and positive (PSP) human serum pools served as controls. The inhibition of the binding of the RBD to hACE2 was calculated as the reduction of the NLuc signal (%) = (1 - NLuc signal of the sample/NLuc signal of the untreated sample) x 100%.

#### Immunoglobulin capture enzyme immunoassay

Immunoglobulin class-specific detection of antibodies was performed by in-house enzyme immunoassays (EIA) (**Fig. 2B**). To this end, black high-binding strip plates (Greiner Bio-One) were coated with 400 ng/well of anti-human IgG (GtxHu-004-J), anti-human IgA (GtxHu-001-G) and anti-human IgM (GtxHu-008-G) goat F(ab’)_2_ fragments (Dianova, Hamburg, Germany), respectively. The procedure was carried out at 4°C for 16 h in 100 µL bicarbonate buffer (pH 9.6). After washing four times with 300 µL wash buffer (TBS pH 7.4 containing 0.05% Tween 20, TBS-T), 300 µL blocking buffer (TBS-T/10% fetal calf serum (FCS)) was added at 37°C for 90 min. After washing again four times, 100 µL of each serum sample were incubated on the sealed plate at a dilution of 1:100 in sample buffer (TBS-T/10% FCS) for 1 h at 37°C. Controls consisting of 100 µL TBS-T/10% FCS without serum as well as NSP and PSP were included. Next, plates were washed again four times with wash buffer and incubated with the NLuc-labelled antigens. The antigens were added at a concentration of 5×10^6^ rlu/100 µL in a total volume of 100 µL sample buffer for 1 h at 37°C. After washing four times, the luciferase activity of NLuc-RBD proteins bound to the plate was quantified by adding 100 µL of NanoGlo reagent. Results were expressed as activity (rlu) after subtracting the blank without serum addition. Cut-off values for all in-house assays were determined through receiver operating characteristic (ROC) curves using GraphPad Prism 10.0 (La Jolla, CA, USA).

## RESULTS

### Routine serologic assessment of serum samples

All 124 participants of the vaccination study underwent routine serology to test for SARS-CoV-2-specific antibodies immediately prior to their first vaccination. At t0, before the first vaccination, sera from 53 participants were non-reactive in the commercial anti-RBD IgG CMIA, anti-N IgG CMIA and line blot. These participants were also available for antibody testing 4 weeks after the first vaccination (t1) and 4 weeks after the second vaccination (t2), and their sera were included in the study. Additional sera from a subset of these vaccinated individuals were gathered 3 months (t3, 42 sera) and 6 months (t4, 26 sera) post-second vaccination, and 4 weeks after the third vaccination (t5, 12 sera). The quantitative anti-RBD-IgG CMIA revealed that all serum samples collected from t1 to t5 were reactive (**Fig. 3**).

**Figure 3:**
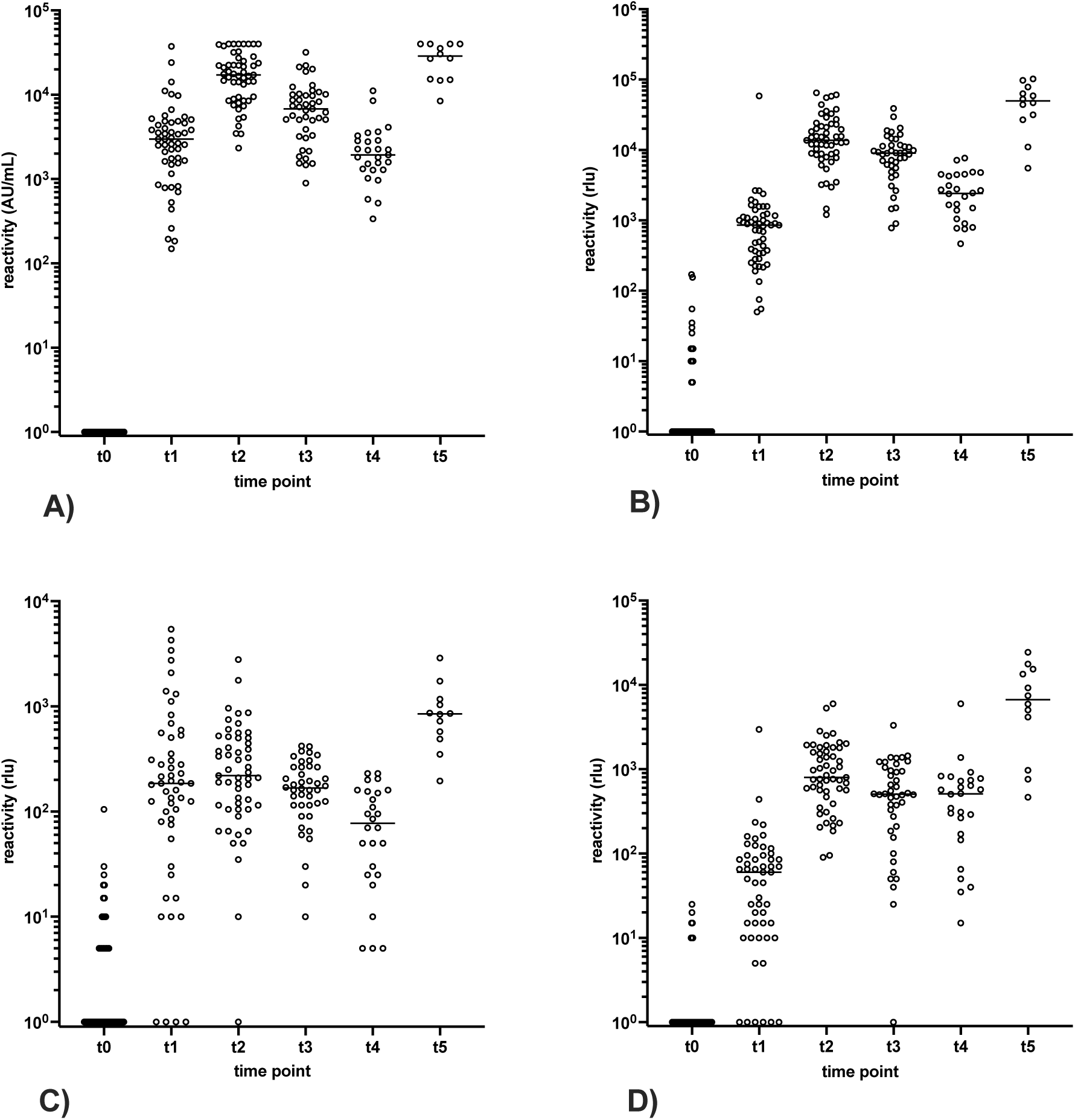
Time kinetics of RBD-specific antibody response in vaccinee sera by *SARS-CoV-2 IgG II Quant* RBD CMIA (Abbott) **(A)** γ-capture EIA **(B)**, µ-capture EIA **(C)**, and α-capture EIA **(D)** at time points t0-t2 (53 sera, respectively), t3 (42 sera), t4 (26 sera) and t5 (12 sera). Horizontal bars delineate the median. Values <0 were set to 1 to allow for logarithmic representation of data.

### Recognition of NanoLuc-tagged S-fragments by serum antibodies

As an initial experimental step, the suitability of NLuc-tagged S fragments containing the RBD derived from the original Wuhan-Hu-1 sequence as diagnostic antigens (**Table 1**) was evaluated. The gamma-chain capture EIA was used to assess the reactivity of constructs comprising the RBD of S (aa 319 to 541), and fragments of the external domain of S spanning from aa 15 to 541, aa 15 to 680, and aa 15 to 1213, respectively, with sera collected at time points t0, t1 and t2 (**Suppl. Fig. 1**). For comparison, an NLuc-labelled NTD fragment (aa 15 to 307) was included. Unprocessed cell culture supernatants adjusted to an NLuc activity of 5×10^6^ rlu per well were used as antigen preparations (rf. Materials and Methods). The reactivity of sera in the quantitative anti-RBD-IgG CMIA (Abbott) served as the gold standard. ROC curves were applied to characterize the diagnostic performance of constructs and to determine cut-off values that allowed discrimination of sera obtained prior to (t0) and after the first vaccination (t1) with 100% specificity. The signal intensity of antibody reactivity measured in the gamma-capture EIA at time points t1 and t2 served as a second criterion to identify the most appropriate NLuc-tagged S protein fragments.

This approach revealed that, amongst the constructs tested, the RBD-containing construct secNLuc-RBD exhibited the highest sensitivity at time point t1 with 92.5% (49/53 sera reactive) (**Suppl. Fig. 1, 2; Table 2**). In comparison, construct secNLuc-S15-541 showed a similar, slightly lower sensitivity of 86.8%, while the sensitivity achieved with constructs secNLuc-S15-680 and secNLuc-spike-CSdel-PPmut at t1 was lower, i.e., at 73.6% and 71.7%, respectively. The secNLuc-NTD construct, which was tested for comparison, displayed a sensitivity comparable to secNLuc-S15-541, i.e., 86.8%. At time point t2, all constructs exhibited a sensitivity of 100% in the gamma-chain capture EIA. In terms of signal strength at time point t2, constructs secNLuc-RBD and secNLuc-S15-541 showed the strongest reactivity, while the reactivity of secNLuc-NTD, secNLuc-S15-680 and secNLuc-spike-CSdel-PPmut in the gamma-chain capture EIA was lower (**Suppl. Fig. 1, 2**).

**Table 2:** Reactivity of secreted NLuc-tagged S antigens in heavy-chain capture EIA. ^1,2^The cut-off to achieve 100% specificity at time point t0 and the ability to distinguish between time points t0 (immediately prior to vaccination) and t1 (four weeks after the first vaccination) was defined by ROC curve analysis and calculation of AUC values.

| Construct | Antibody class | Cut-off<sup>1</sup> (rlu) | AUC t0/t1<sup>2</sup> | Sensitivity t1 | Sensitivity t2 |
| --- | --- | --- | --- | --- | --- |
| secNLuc-RBD | IgG | ≥180 | 0.9966 | 92.5% (49/53) | 100% (53/53) |
| secNLuc-RBD | IgM | ≥115 | 0.8984 | 66% (35/53) | 75% (40/53) |
| secNLuc-RBD | IgA | ≥30 | 0.9532 | 58.5 (31/53) | 100% (53/53) |
| secNLuc-S15-541 | IgG | ≥90 | 0.9845 | 86.8% (46/53) | 100% (53/53) |
| secNLuc-S15-680 | IgG | ≥50 | 0.9591 | 73.6% (39/53) | 100% (53/53) |
| secNLuc-spike-CSdel-PPmut | IgG | ≥350 | 0.9778 | 71.7% (38/53) | 100% (53/53) |
| secNLuc-NTD | IgG | ≥90 | 0.9797 | 86.8% (46/53) | 100% (53/53) |

These experiments demonstrated that the secNLuc-RBD antigen was most sensitively recognized by human sera after the first vaccination, allowed for the best differentiation between sera obtained before and after the first vaccination, and yielded the strongest signals after the second vaccination. Therefore, the construct was chosen for further experimentation.

As the sVNT does not distinguish between antibody classes, all serum samples collected at time points t0 to t5 were analysed using EIA to determine their antibody class-specific reactivity. Cut-off values for IgA- and IgM-EIA were identified through ROC curve analysis (**Suppl. Fig. 2**). This approach revealed that secNLuc-RBD reacted most strongly with IgG antibodies from t1 to t5 (**Fig. 3**). The IgG reactivity pattern in the gamma-chain capture EIA was almost indistinguishable from that found by the commercial anti-RBD CMIA. A notable increase was evident from t1 to t2, with the highest levels reached at time points t2 and t5. In contrast, IgG levels decreased from t2 to t3 to t4.

The reactivity pattern of IgA with secNLuc-RBD highly resembled that of IgG, but tended to be elevated after the third vaccination (t5), and sustained a minor descent from t2 to t4. Overall, the signal strength of IgA was approximately 10-fold lower as compared to that of IgG. Dissimilar to IgG and IgA, the maximum IgM levels were detected at t1 and t5, while no increase occurred from t1 to t2. Between t2 and t4, RBD-specific IgM antibodies decreased more markedly than IgG and IgA antibodies. Additionally, IgM antibody levels were significantly lower than those of IgG.

### Detection of inhibitory antibodies by sVNT

Employing the secNLuc-RBD construct as a diagnostic antigen in our in-house sVNT showed the effectiveness of this experimental approach for detecting antibodies that impede RBD binding to hACE2 (**Fig. 4**) To ensure 100% specificity when discriminating between sera obtained at time points t0 and t1, the cut-off was set at 25% inhibition (**Suppl. Fig. 2**), resulting in a sensitivity of 98% at t1 (**Table 2**). RBD-specific antibody detection by the in-house sVNT at t2 was achieved with a sensitivity and specificity of 100%. At time points t3, t4 and t5, 42 out of 42 sera, 24 out of 26 sera and 12 out of 12 sera were reactive in the in-house sVNT.

**Figure 4:**
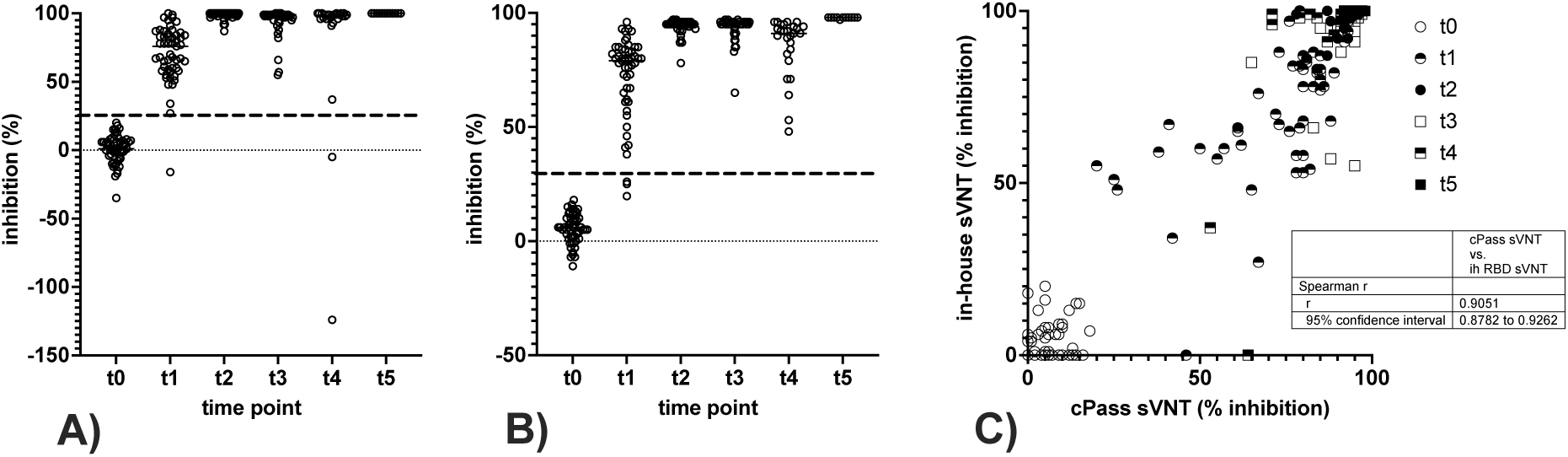
Detection of inhibitory antibodies by sVNT. **(A)** In-house sVNT employing the secNLuc-RBD construct was used to determine antibodies inhibiting binding of the RBD to hACE2 in vaccinee sera at time points t0-t5. The cut-off determined by ROC curve analysis (25% inhibition) is indicated by the dashed line. Horizontal bars delineate the variable median. **(B)** Analysis of vaccinee sera by the cPass sVNT at time points t0-t5. The cut-off value according to the manufactureŕs manual is indicated by the dashed line (30% inhibition), and horizontal bars delineate the variable median. **(C)** Correlation of in-house sVNT and cPass sVNT, values <0% were set to 0%, and the correlation coefficient was calculated according to Spearman. Ih: in-house.

Compared to the commercial cPass sVNT (**Fig. 4**), the in-house sVNT displayed comparable or slightly improved sensitivity at time point t1 (52 out of 53 sera were reactive, with 98% sensitivity vs. 94%) (**Table 3**). One serum sample non-reactive at t1 in the in-house sVNT was reactive in the cPass sVNT and in the heavy-chain capture EIA, however, exhibited low IgG levels and moderate IgM levels and was IgA negative. All three serum samples that did not react in the cPass assay at t1 also were nonreactive in IgG, IgA and IgM capture EIA.

**Table 3:**
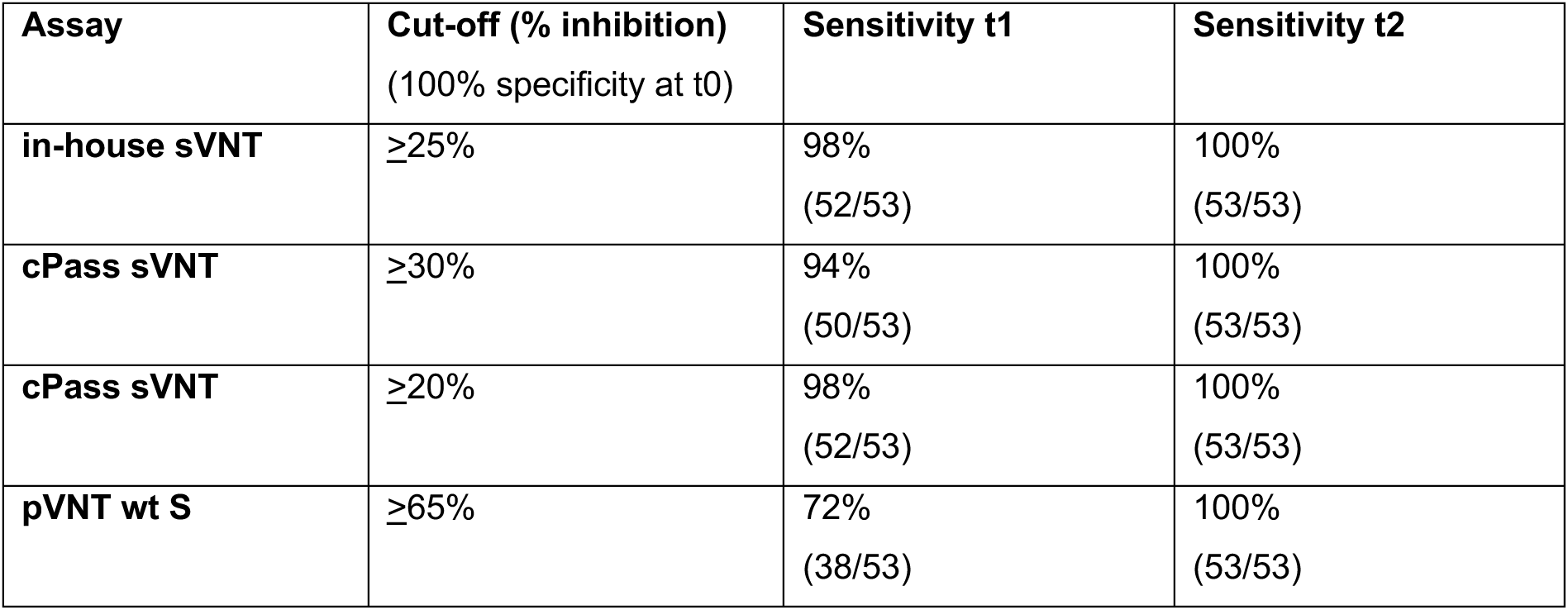
Sensitivity of sVNT and pVNT in sera obtained at t0 to t2.

At time point t4, the sensitivity of the in-house sVNT was slightly lower as compared to cPass. Both serum samples that were non-reactive in the in-house sVNT at t4 exhibited reactivity in cPass and in the IgG EIA, had low levels in the IgA EIA, and were negative in the IgM EIA. At time points t2 and t5, all sera displayed reactivity in both sVNTs. ROC curve analysis of the cPass sVNT results revealed a 98% sensitivity and specificity in discriminating sera at t0 and t1 using a cut-off of 20% inhibition, which differs from the specifications of the CE/IVD certified product by the manufacturer (**Table 3, Suppl. Fig. 2**).

While the Abbott CMIA and the capture EIAs demonstrated a considerable decrease in antibody levels following the second vaccination, the cPass and in-house sVNT analysis indicated a much less significant decline. Both sVNTs produced consistent results at time points t0 and t5, but varied in signal strength more widely at t1 to t4. The greatest disparities were observed at time points t3 and t4 (**Fig. 4**). Overall, the correlation between both sVNTs was strong, as indicated by the Spearman r value of 0.91 (95% confidence interval: 0.88 to 0.93).

To evaluate the functional significance of inhibitory antibodies by sVNT, the formation of neutralizing antibodies was examined via VSV-pVNT (**Fig. 5**). Employing ROC curve analysis and vaccinated versus non-vaccinated differentiation as criteria, we established a cut-off of 65% inhibition for detection of neutralising antibodies at time point t1 which ensured 100% specificity (**Suppl. Fig. 2**). This resulted in 72% sensitivity of the pVNT at time point t1 (**Table 3**). At t2, all human sera were reactive in the pVNT. Differences in outcomes between the pVNT, the in-house sVNT and the cPass sVNT were mainly noticeable in sera collected at t1. Taken together, a strong correlation between the results of the in-house sVNT and pVNT was observed with Spearman r of 0.9 (95% confidence interval: 0.86 to 0.92) (**Fig. 5**).

**Figure 5:**
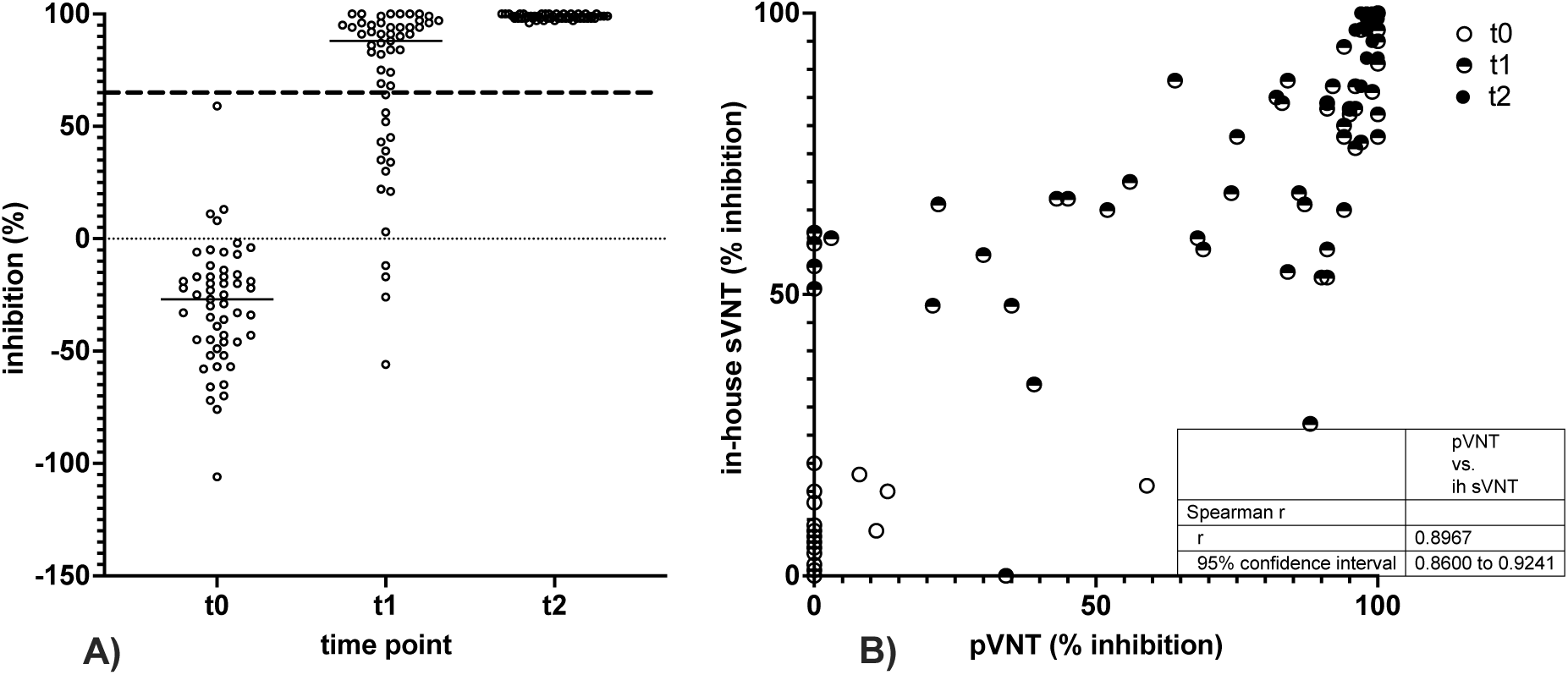
VSV-pseudotyped pVNT, **(A)** neutralising activity of sera obtained from 53 vaccinees on t0, t1 and t2 in pVNT, dashed line depicts cut-off, horizontal lines show the variable median. **(B)** comparison of results obtained in in-house sVNT and pVNT in sera obtained on t0, t1, and t2. Values <0 were set to 0, and the correlation coefficient was calculated according to Spearman.

Using the pVNT as the gold standard, the effectiveness of sVNTs, Anti-RBD CMIA and Anti-RBD IgG EIA in predicting neutralising antibodies among the 159 sera collected from t0 to t2 was analysed. By raising the cut-off of the in-house sVNT to over 73% inhibition, the detection of neutralising antibodies by pVNT could be predicted with a 98.6% specificity and 87.8% sensitivity. At a cut-off of over 76.5% inhibition, the commercial cPass sVNT showed 100% specificity and 95.6% sensitivity (**Table 4**). By increasing the cut-off to over 1706 AU/mL in the Anti-RBD CMIA, 95.65% specificity and 96.67% sensitivity were achieved relative to the pVNT benchmark. For the in-house RBD IgG EIA, a cut-off of over 1103 rlu resulted in 100% specificity and 75.6% sensitivity. Adjusting the cut-off of the in-house RBD IgG EIA to over 617.5 rlu enabled the prediction of neutralising antibodies with 97.1% specificity and 92.2% sensitivity (**Table 4**). As expected, raising the cut-off values caused a significant decline in the sensitivity of all assays regarding differentiation between vaccinated and unvaccinated individuals. At time points t0 and t1, the in-house sVNT, cPass sVNT, Anti-RBD CMIA and Anti-RBD IgG EIA showed sensitivity reductions to 51.9%, 60.4%, 69.8% and 60.4%, respectively.

**Table 4:**
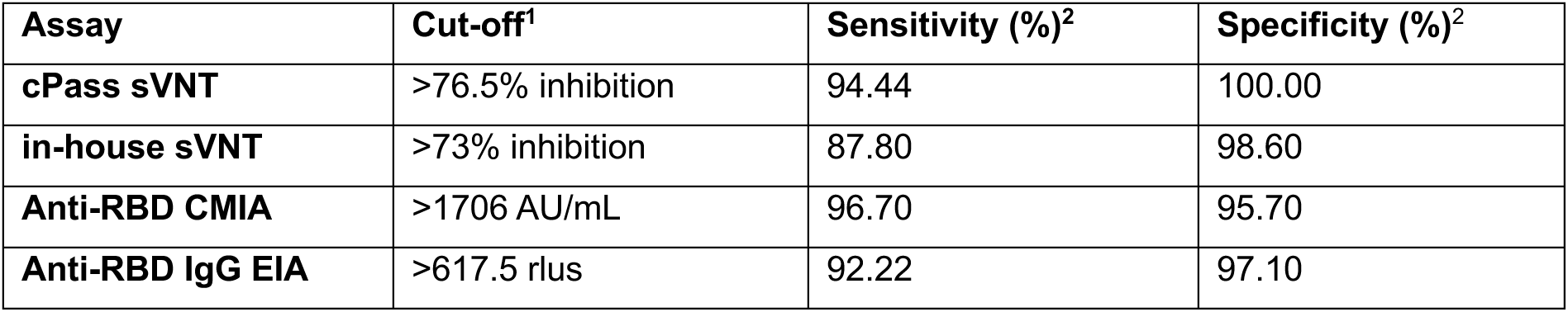
Prediction of reactivity in pVNT by sVNT, CMIA and EIA. ^1^ optimal threshold values were defined by ROC curve analysis of data and calculation of Youden’s index. ^2^ as compared to pVNT as the gold standard in sera obtained from t0 to t2.

### Detection of inhibitory antibodies reactive with the Omicron BA.1 variant

Finally, we investigated whether the RBD-based in-house sVNT would detect the induction of cross-inhibitory antibodies against Omicron BA.1 upon vaccination with wt S. To this end, inhibitory antibodies against the wt (in-house sVNT) and Omicron BA.1 RBD (in-house BA.1 sVNT), respectively, were measured in nine vaccinees whose serum samples were taken at all time points of the vaccination study. The determination of neutralising antibodies against wt S and BA.1 S via in-house pVNT served as a control.

The pVNT results indicated that all vaccinees tested required three vaccinations (t5) for the induction of a robust cross-neutralizing antibody response against Omicron BA.1 S (**Fig. 6**). At earlier time points, none (t1), four (t2), three (t3) and a single (t4) out of nine serum samples contained neutralising antibodies against Omicron BA.1 S with an inhibitory activity at least 65% at a serum dilution of 1:20. In contrast, neutralizing antibodies against wt S were detected in all sera obtained from t2 to t5, as well as in six out of nine sera gathered at t1 (**Fig. 6**). Significantly higher neutralising antibody levels against wt S were observed from t1 to t4 (**Fig. 6**). At t5, all sera neutralized wt S and BA.1 S with nearly 100% activity at a dilution of 1:20.

**Figure 6:**
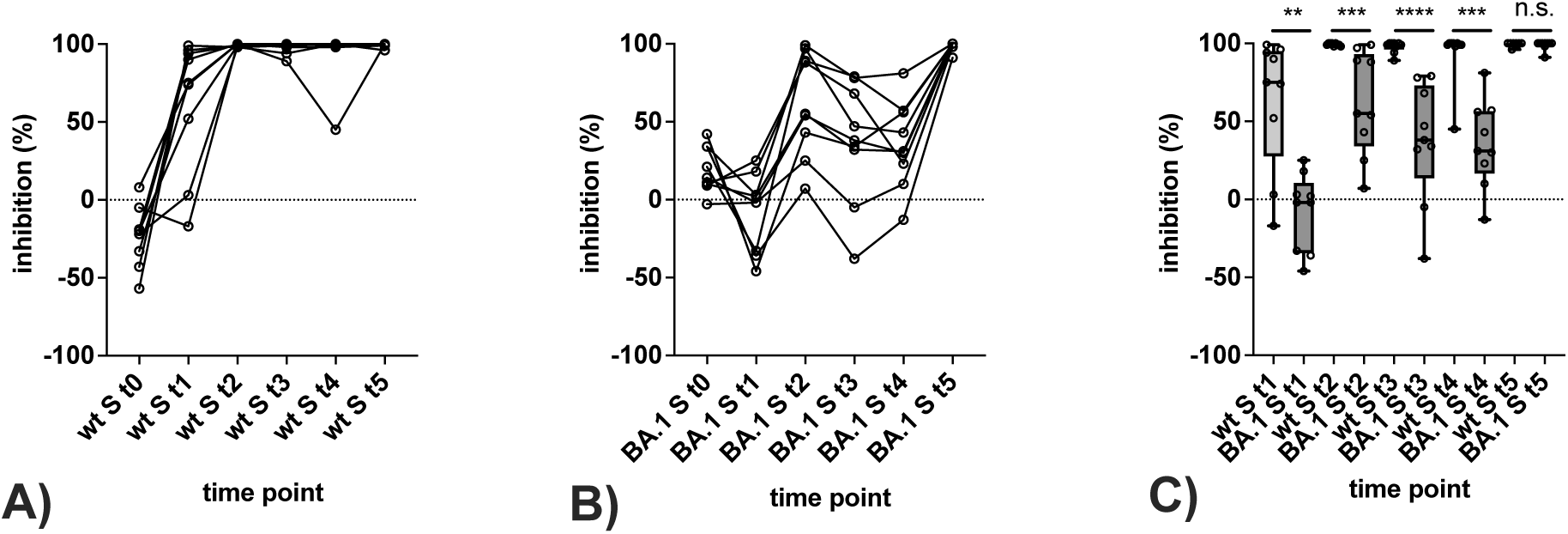
Detection of antibodies reactive with the Omicron BA.1 variant in pVNT. **(A, B)** Consecutive sera of nine vaccinees obtained at time points t0 to t5 were tested for neutralising antibodies against wt S (A) and BA.1 S (B) by sVNT. **(C)** Differences in reactivity with the wt S and BA.1 S at time points t0 to t5. Horizontal lines represent the variable median. The significance of differences in reactivity at individual time points was determined by the Mann-Whitney test (P >0.05: n.s., P <0.05: *, P <0.01: **, P <0.001: ***, P <0.0001: ****).

Overall, the pattern of antibody reactivity in the in-house sVNTs correlated with the outcome of the pVNT in the detection of BA.1-reactive neutralizing antibodies (**Fig. 7**). At time point t0, a higher level of background reactivity of sera with the Omicron BA.1 RBD as compared to the wt RBD was observed. Raising the cut-off of the in-house BA.1 sVNT to 50% inhibition to avoid false-positive results at t0 led to equivalent differentiation between Omicron BA.1 reactive and non-reactive samples in the sVNT and pVNT. As observed by pVNT, none of the sera reacted with Omicron BA.1 at t1. Three wt S-based vaccinations were necessary to elicit a robust reactivity in the in-house BA.1 sVNT in all sera. Levels of inhibitory antibodies against the wt RBD were significantly higher in all samples from t1 to t5 (**Fig. 7**).

**Figure 7:**
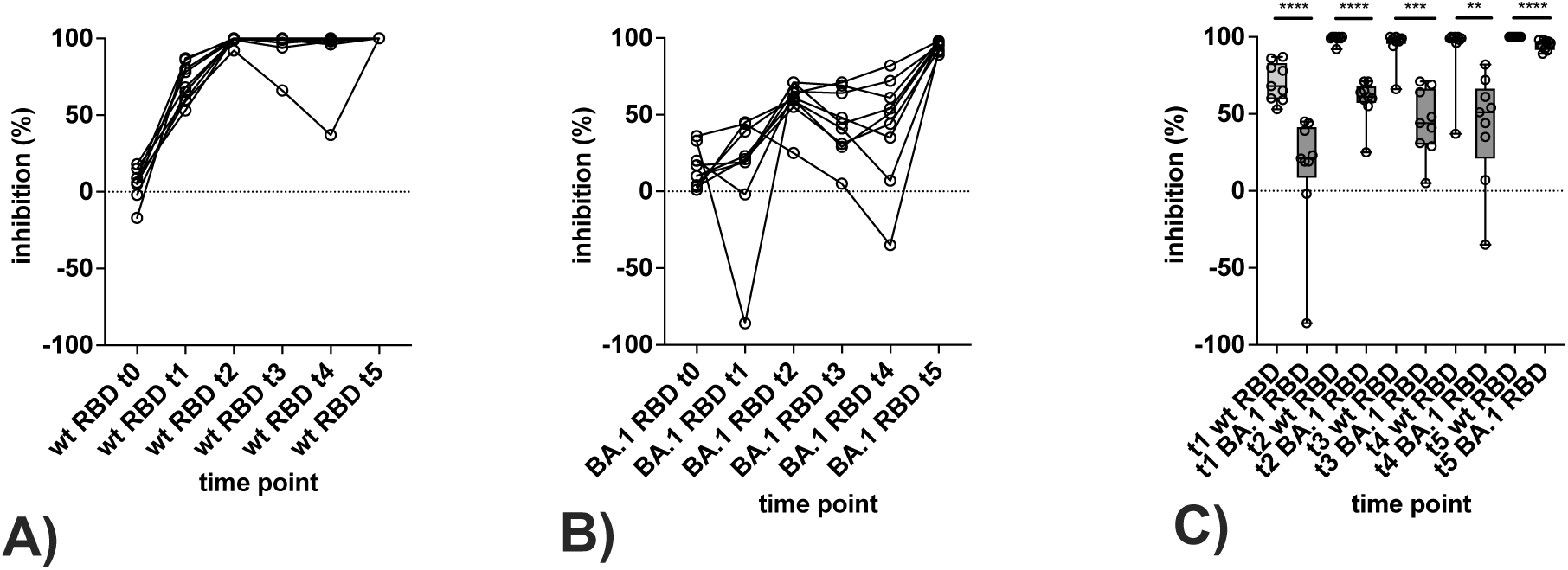
Detection of antibodies reactive with the Omicron BA.1 variant in in-house sVNT. **(A, B)** Consecutive sera of nine vaccinees obtained at time points t0 to t5 were tested for antibodies reactive with wt RBD (A) and BA.1 RBD (B) by sVNT. **(C)** Differences in reactivity with the wt RBD and BA.1 RBD at time points t0 to t5. Horizontal lines represent the variable median. The significance of differences in reactivity at individual time points was determined by the Mann-Whitney test (P >0.05: n.s., P <0.05: *, P <0.01: **, P <0.001: ***, P <0.0001: ****).

In contrast, the determination of variant-reactive antibodies by heavy chain capture EIA for comparison revealed that this method less reliably distinguished between wt and Omicron BA.1-reactive antibodies, despite using the same antigens as in the in-house sVNTs (**Suppl. Fig. 3**). Significant differences in reactivity were observed in the IgG EIA at time points t3 and t4, in IgA EIA at t2 and t3, and in IgM EIA at t3 and t4, respectively. The relative differences in reactivity with the wt RBD and Omicron BA.1 RBD, however, were less pronounced as compared to sVNT and pVNT.

## DISCUSSION

In summary, the utilisation of NLuc-labelled S protein fragments in in-house sVNT and EIA permit comprehensive characterisation of antibodies induced by SARS-CoV-2 infection and/or vaccination. Both assay systems are characterised by high specificity, sensitivity and reproducibility and can be conveniently performed in a routine laboratory environment. As the in-house assays employ unpurified cell-culture supernatants as the source of diagnostic antigens, they are easy to conduct.

NLuc-tagged SARS-CoV-2 S protein fragments have been demonstrated to be effective as diagnostic antigens in liquid-phase luciferase immune precipitation (LIPS) assays, particularly when attaching the tag to the N-termini of proteins [35]. In preliminary experiments, we confirmed the superiority of N-terminally over C-terminally tagged RBD constructs for our in-house sVNT, which prompted us to choose N-terminal labelling for all diagnostic antigens (data not presented). While we cannot rule out that longer S fragments or the NTD might have yielded better results when labelled C-terminally, we opted not to investigate this hypothesis further and focussed on the RBD instead. As infections with Omicron and its successors have decreased the diagnostic sensitivity of antibody assays based on the ancestral Wuhan strain of SARS-CoV-2, the choice of antigens has to be reconsidered [34]. Thus, we established an assay system that enables rapid adaptation to emerging viral strains. As the shortest well-performing diagnostic antigen currently available, the RBD offers an advantageous option when studying immune responses to variants such as Omicron, which harbour more than 30 mutations in the S sequence [23].

In addition to IgG1, IgM and, to a lesser extent, IgA antibodies have been identified as crucial components of neutralising antibody responses [38]. This prompted us to develop a capture EIA to investigate isotype-specific responses to viral variants. Implementing the NLuc-tagged wt RBD, the in-house EIA clearly discriminated between IgM, IgA and IgG reactivity. However, when testing vaccinee sera using an NLuc-tagged Omicron BA.1 RBD as the diagnostic antigen, the EIA exhibited inferior performance in discriminating between variant-specific IgG antibodies compared to the sVNT and pVNT.

In contrast to EIA-based serologic assays, the sVNT detects antibodies of all isotypes, which may result in higher sensitivity and better negative predictive values compared to isotype-specific tests, as shown for the cPass sVNT versus commercial IgG enzyme immunoassays [27]. We verified this observation when comparing the in-house capture EIA and sVNT using the NLuc-tagged RBD as the antigen. However, it has been reported that the cPass sVNT may overestimate low neutralising antibody levels when using the cut-off value of 20% recommended by its manufacturer for the RUO (research use only) version upon release, requiring a threshold of up to 50% when analysing high-titre sera [28]. Subsequently, this issue was resolved by raising the cut-off value to 30% when launching the CE/IVD certified cPass sVNT on the market [25], at a slight expense of sensitivity while increasing specificity [30, 32]. Alternatively, a threshold area between 15% and 35% indicating equivocal results could be defined, which would warrant confirmation by other VNTs, especially when testing low-titre sera with a focus on individual results [32].

Neutralising antibody titres exceeding the quantifiable range of the assay, which are often found in vaccinees, may require adjustment of the cut-off values. It has been recommended that ROC curves be calculated [31] or end-point dilution of sera be performed. We set the cut-off value for the in-house sVNT to 25% to avoid false-positive results in sera diluted 1:20. As the range of NLuc signals surpasses that of HRP, the reporter enzyme deployed in the cPass sVNT, displaying remaining hACE2 binding capacity instead of inhibition could improve resolution when testing high-titre sera. In the case of investigating variant-specific antibodies, cut-off values may have to be adapted accordingly. A cut-off value of 65% was established for optimal discrimination between Omicron BA.1 reactivity vs. non-reactivity in the in-house sVNT.

Using the reactivity of sera in the pVNT as the benchmark, the cut-off of the sVNT needs adjusting to improve agreement between the assays. Our data show that a cut-off of over 75% inhibition in the sVNT enables prediction of the presence of neutralising antibodies in sera with high specificity and reasonable sensitivity. Hence, the same method can determine the appropriate cut-off titres for the anti-RBD CMIA and anti-RBD IgG EIA, respectively. With an extended time interval from vaccination or infection, determination of serum antibody levels with non-functional tests such as CMIA or EIA could pose problems due to antibody waning. Contrary to the commercial CMIA and the in-house EIA, which both revealed a decline in antibody levels in sera collected from t2 to t4, the inhibitory capacities of antibodies determined by in-house pVNT and sVNT persisted at high levels, suggesting affinity maturation. This reinforces the requirement to complement routine antibody quantification with functional characterisation of the humoral immune response. Unlike the in-house pVNT that involves the entire ectodomain of the S protein, the in-house sVNT solely measures antibodies that interfere with the RBD-hACE2 interaction, which pertains to most neutralising antibodies; however, antibodies that bind to different epitopes will escape detection [29].

## Conclusion

To conclude, our data show that the in-house sVNT and, to a lesser degree, the EIA utilising the NLuc-tagged RBD enable rapid quantification and functional evaluation of variant-specific antibodies in a hybrid-immune population that is prevalent worldwide these days. Conceivable applications comprise monitoring the efficacy of antibody-based therapy vaccines, screening drug candidates that interfere with the RBD-hACE2 binding, and following up on prolonged COVID-19 courses in high-risk groups in clinical settings. As highly flexible modular systems, the tests presented in this study can readily be adapted to further emerging virus variants.

## Data Availability

All data produced in the present study that are not contained in the manuscript are available upon reasonable request to the authors.

## ACKNOWLEDGEMENTS

The plasmid pCG1-SARS-2-S was kindly provided by Prof. Dr. Stefan Pöhlmann (Infection Biology Unit, German Primate Centre, Göttingen, Germany). We are grateful for the donation of plasmid pCAspike-CSdeleted-PPmutation to Prof. Dr. Florian Krammer (Department of Microbiology, Icahn School of Medicine at Mount Sinai, New York, NY, USA).

Parts of this study regarding the methodological establishment of the in-house sVNT and EIA were submitted in 2023 by TG to the Medical Faculty of the University of Münster, Münster, Germany, as their doctoral thesis entitled “Etablierung serologischer Nachweisverfahren zur Varianten-spezifischen Detektion und Differenzierung von Antikörpern gegen SARS-CoV-2”. We thank Sabine Lima, Klaudia Swierczek and Maik Voskort for their excellent technical support.

This study was partially funded by the Ministry of Labour, Health, and Social Affairs of the state of North Rhine-Westphalia, Germany (grant CPS-1-1G). The Institute of Virology is part of the Virus Alliance North Rhine-Westphalia (VIRAL.NRW), which is supported by the Ministry of Culture and Science, North Rhine-Westphalia, Germany.

---

**Supplemental Figure 1:**
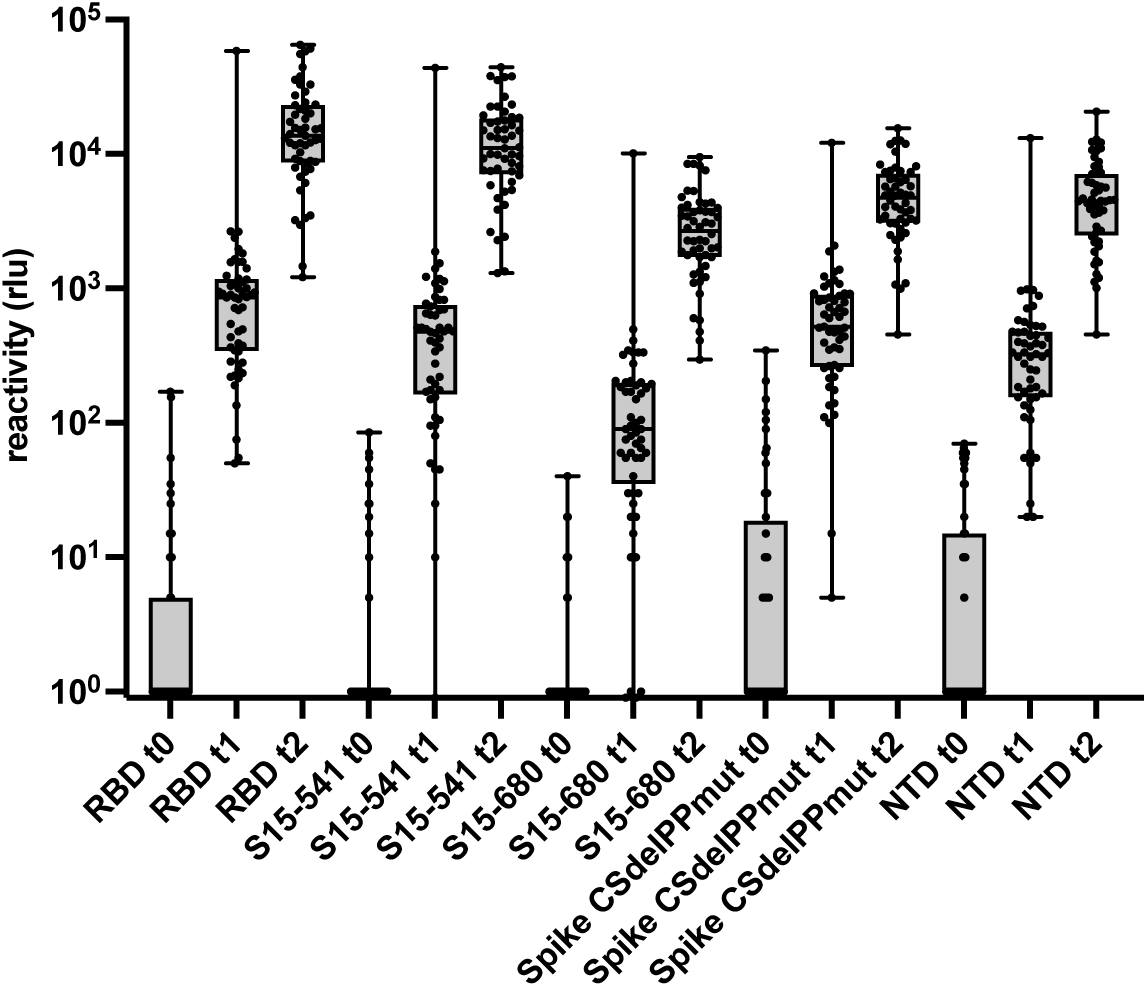
Recognition of NLuc-tagged S protein constructs by human sera. Reactivity of constructs secNLuc-RBD (RBD), secNLuc-S15-541 (S15-541), secNLuc-15-680 (S15-680), secNLuc-spike-CSdel-PPmut (Spike CSdelPPmut) and secNLuc-NTD (NTD) was quantitated by gamma-chain capture EIA as described in Materials and Methods. 53 consecutive sera of vaccinees obtained immediately before vaccination (t0, open circles), four weeks after the first vaccination (t1, grey circles) and four weeks after the second vaccination (black circles) were tested at a dilution of 1:100. Reactivity is given as rlu. Values <0 were set to 1.

**Supplemental Figure 2:**
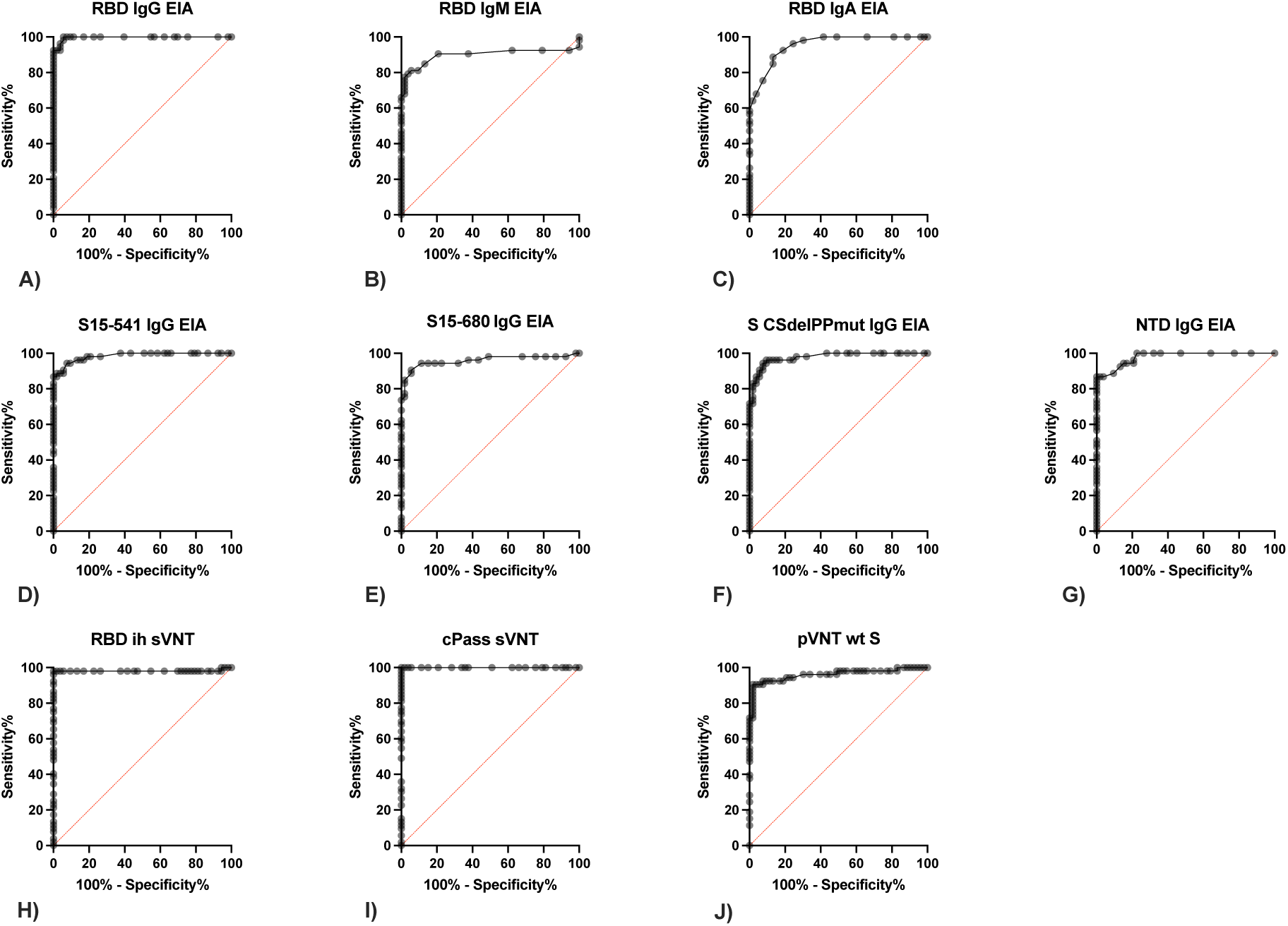
Diagnostic performance of antigen constructs. Discrimination of serum samples obtained at t0 and t1 was determined by calculating ROC curves. **(A-C)** Reactivity of sera (n=53) with secNLuc-RBD in γ-capture EIA (A), µ-capture EIA (B) and α-capture EIA (C). **(D-G)** Reactivity in gamma-capture EIA with secNLuc-15-541 (D), secNLuc-15-680 (E), secNLuc-spike-CSdel-PPmut (F) and secNLuc-NTD (G)**. (H-J)** reactivity of sera in in-house sVNT with secNLuc-RBD (H), commercial sVNT (cPass) (I), and pVNT with wt S (J).

**Supplemental Figure 3:**
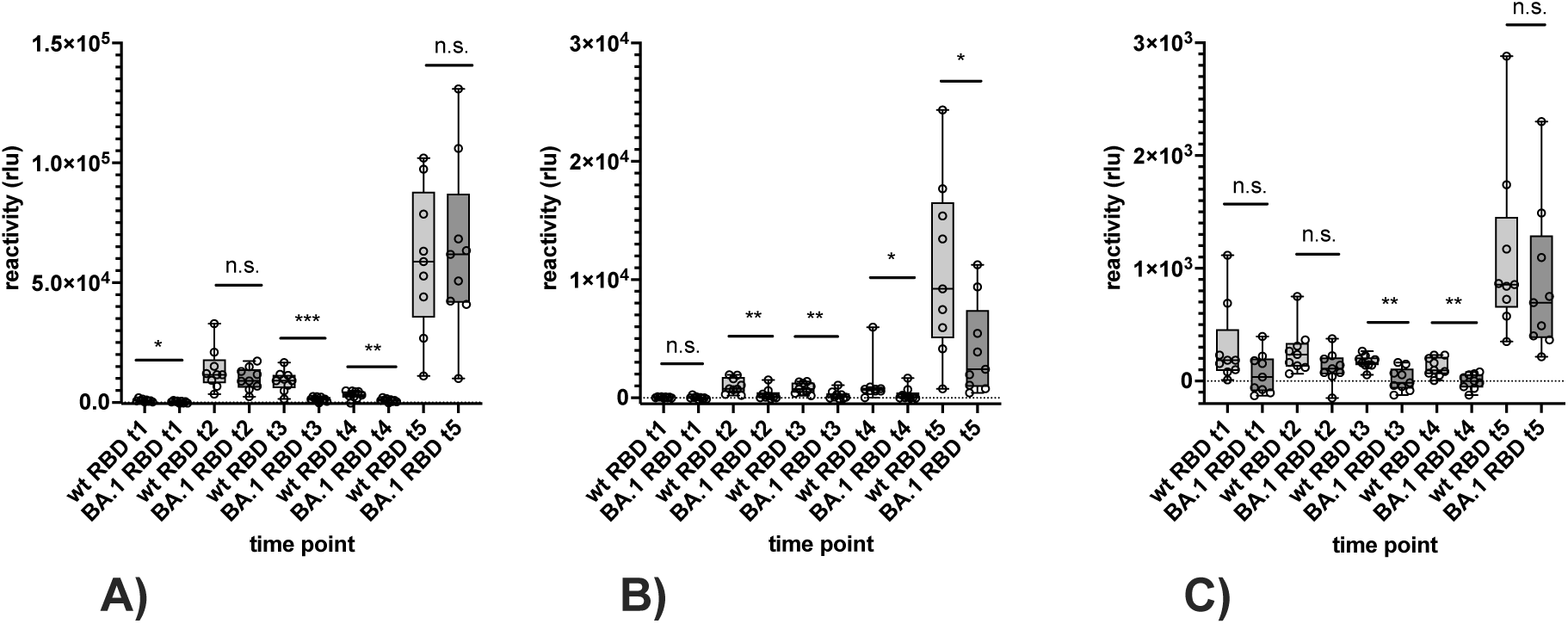
Detection of antibodies reactive with the Omicron BA.1 variant in IgG EIA **(A)**, IgA EIA **(B)** and IgM EIA **(C)**. Horizontal lines represent the variable median. The significance of differences in reactivity at time points t1 to t5 was determined by the Mann-Whitney test (P >0.05: n.s., P <0.05: *, P <0.01: **, P <0.001: ***, P <0.0001: ****).

## Notes

### Competing Interest Statement

The authors have declared no competing interest.

### Author Declarations

Ethik-Kommission der Aerztekammer Westfalen-Lippe und der Westfaelischen Wilhelms-Universitaet, Muenster, Germany, gave ethical approval for this work (2021-039-f-S).

